# Technical Development and Implementation of 3D-QALAS on a 1.5T MR-Linac for the Brain: A Prospective R-IDEAL Stage 0/1 Technology Development Report

**DOI:** 10.64898/2026.03.09.26347967

**Authors:** Lucas McCullum, Ashley Harrington, Brian A. Taylor, Ken-Pin Hwang, Clifton D. Fuller

## Abstract

**Background and Purpose:** Quantitative relaxometry on the integrated MRI / linear accelerator (MR-Linac) at high isotropic resolution is currently limited due to prohibitively long scan times and limited field-of-views. Therefore, the purpose of this study was to assess the technical feasibility of the 3D-QALAS technique on the 1.5T MR-Linac which has the ability to acquire whole-brain 1 mm isotropic quantitative T1, T2, and PD maps along with multiple synthetic images in a 7 minute acquisition time.

**Materials and Methods:** A 1 mm isotropic 3D-QALAS acquisition was scanned in both phantoms and a healthy volunteer on the 1.5T Elekta Unity MR-Linac device with scan times around seven minutes. A test-retest protocol across five independent sessions for the phantom was conducted. The correlation, repeatability, and reproducibility between measured and reference quantitative T1, T2, and PD values were determined in the phantom. Distortion was also studied. Vendor-provided reconstruction through SyMRI was performed to extract synthetic images and brain volume metric assessments on a healthy volunteer.

**Results:** The slope and concordance between the measured and phantom reference values was 1.02 (1.00), 1.09 (0.90), and 0.99 (1.00) for T1, T2, and PD, respectively. Median distortion across the phantom remained below 2 mm. The repeatability and reproducibility coefficient-of-variation (CoV) was under 8% for all measured values. The measured brain volumes in the healthy volunteer was within expected age-adjusted reference values.

**Discussion:** The technical feasibility of using 3D-QALAS on the integrated 1.5T MR-Linac was confirmed. Applying this technique to the head and neck adaptive radiation therapy workflow will provide new opportunities to integrate quantitative imaging relaxometry biomarkers at 1 mm isotropic resolution.

**Graphical Abstract:** 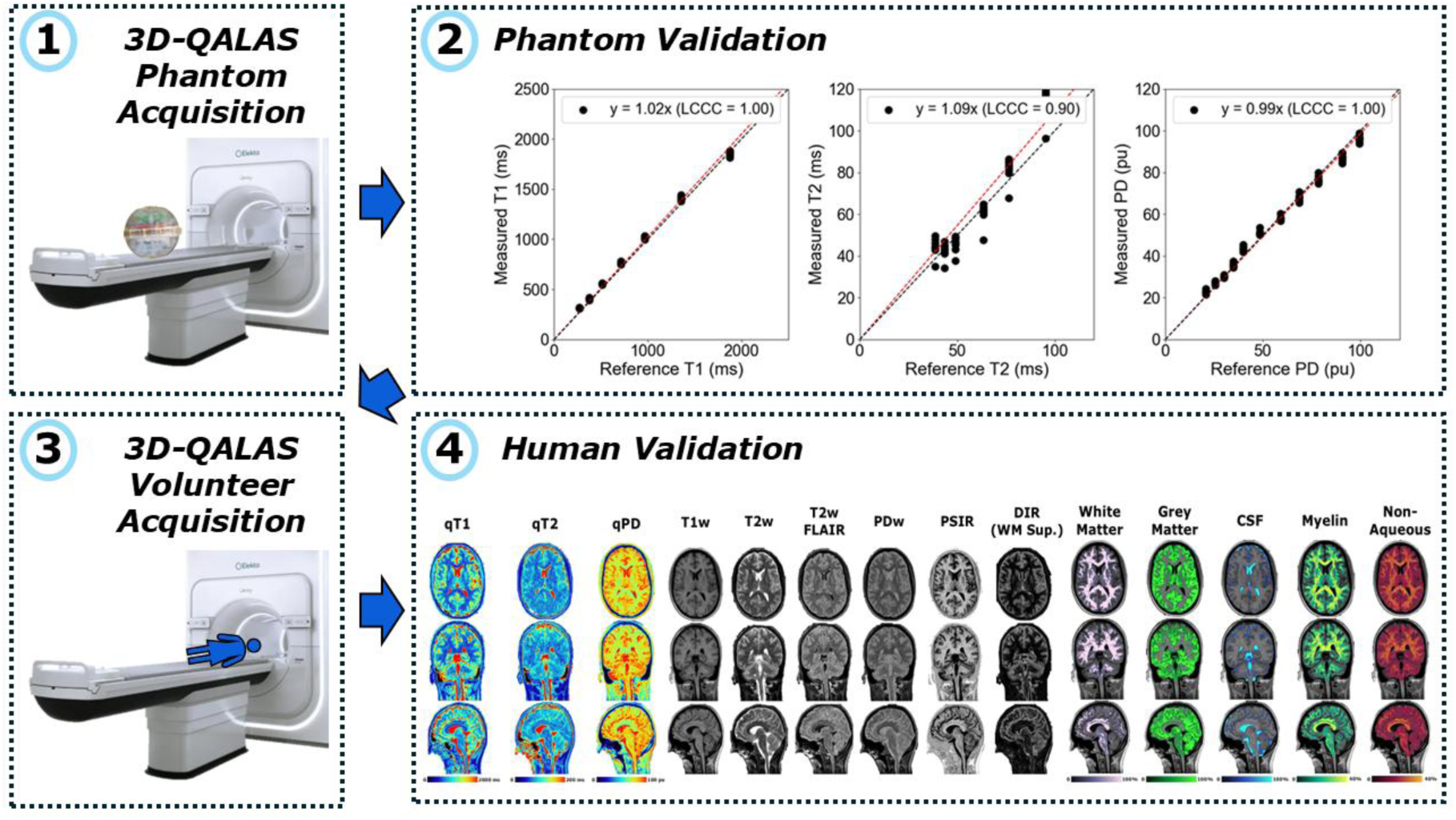

## 1. Introduction

The combined magnetic resonance imaging (MRI) linear accelerator (Linac), or MR-Linac, has seen increasing application in the treatment of cancers of the head and neck^1^. Head and neck cancer is a good candidate for treatment on the MR-Linac due to the required soft tissue contrast^2^, frequent anatomical changes throughout radiation therapy^3^, and close proximity of organs-of-interest (OOIs) to the intended treatment target^4^. The close proximity to neighboring OOIs further warrants high resolution imaging to ensure accurate target localization and safe delivery of radiation^5,6^. On the MR-Linac, the increased spatial resolution also leads to improved contouring accuracy across the dice score, Hausdorff distance, and mean distance to agreement metrics^7^. Even though contouring could be done on just the high-resolution anatomic images, quantitative imaging can be supplemented to improve accuracy^8,9^ and similar spatial resolutions should be used between the two images to reduce spatial blurring^10,11^. Further, acquiring at small voxel sizes will allow for stereotactic body radiation therapy (SBRT) treatments of the head and neck^12,13^. More formerly, the consensus statement from the German Society for Radiation Oncology (DEGRO) and for physics and technology in stereotactic radiotherapy of the German Society for Medical Physics (DGMP) recommends a maximum geometric accuracy of 1 mm for stereotactic radiosurgery^14^ (SRS) and 1.5 mm for SBRT^15^.

Another motivation for high resolution quantitative imaging is for the identification of brain metastases which, in a recent consensus recommendation, should be imaged in at most 1.5 mm resolution on 1.5T and 1 mm resolution on 3T MRI scanners^16^ which was corroborated by another study from Cancer Care Ontario^17^.

In the head and neck, quantitative T1 and T2 mapping has been applied to describe patterns of response to radiation therapy with encouraging results^18^. Although promising, this specific study utilized two separate quantitative mapping techniques (variable flip angle for T1 and multi-echo spin-echo for T2) at a 2 x 2 x 4 mm^3^ voxel resolution, motivating future researchers to improve spatial resolution and develop combined imaging methods for simultaneous T1 and T2 quantification to better characterize tumor dynamics throughout radiation therapy. One possible solution on the MR-Linac is the 2D multi-dynamic multi-echo (MDME) sequence^19^ from SyntheticMR^20^ (Linköping, Sweden), evolved from its original implementation^21^, which can simultaneously quantify T1, T2, and PD along with synthetically-generate contrast-weighted images through the accompanying post-processing software, SyMRI. In a previous study, McCullum et al. studied the technical feasibility of the 2D-MDME sequence on the 1.5T MR-Linac (Unity; Elekta AB; Stockholm, Sweden) and found mean bias in phantoms under 11% and correlations above 0.97 across multiple repeats^22^. In a follow-up study, McCullum et al. optimized the 2D-MDME sequence across varying acquisition parameters and found that a minimum of a 3 mm slice thickness while preserving a 1 mm in-plane resolution is recommended to maintain minimal noise as measured by the coefficient of variation in both phantoms and healthy volunteers^23^. Although promising, a clear gap exists to reduce the slice thickness to the in-plane spatial resolution of 1 mm, however this is not possible using the previously studied 2D-MDME sequence on the MR-Linac. Fortunately, a 3D version of the sequence capable of 1 mm isotropic acquisitions, namely Quantification using an interleaved Look-Locker Acquisition Sequence with T2 preparation pulse (3D-QALAS) / T2-prepared Multiple Inversion Recovery (TPMID) / SyntAc on Philips systems, has been developed^24^. This technique originally showed clinical acceptability in cardiac applications^25,26^, however newer translations have demonstrated its quantitative accuracy and repeatability in the brain in both multi-center^27^ and multi-vendor studies^28^ as well as in the breast^29^ and liver^30^. Further, its acquisition time has been accelerated by integrating compressed sensing reconstruction leading to clinically feasible scan times^31–33^ while improving repeatability^34^ compared to traditional techniques like Driven Equilibrium Single Pulse Observation of T1 and T2 (DESPOT^35^). Another benefit of 3D-QALAS compared to 2D-MDME is its increased robustness to the magnetization transfer effect which reduces the quantitative T1 values by up to 35% in the 2D-MDME sequence^36,37^.

At the time of this writing, no studies to the author’s knowledge have evaluated the technical feasibility of the 3D-QALAS sequence on the MR-Linac device. Therefore, in this study, our goal is to characterize the quantitative accuracy and repeatability of the 3D-QALAS sequence on a 1.5T MR-Linac for longitudinal biomarker study applications in addition to its geometric accuracy for precise targeting and segmentation pipelines in both phantoms and a healthy volunteer. This will be presented using the radiotherapy-predicate studies, idea, development, exploration, assessment, and long-term study (R-IDEAL) framework, as recommended by the MR-Linac Consortium, completing Stage 0 (radiotherapy predicate studies) and Stage 1 (first time use) systematic evaluations^38^.

## 2. Methods and Materials

### 2.1. MRI Acquisition Parameters

To evaluate the performance of the 3D-QALAS sequence, MRI scans were performed on a 1.5T MR-Linac (Unity; Elekta AB; Stockholm, Sweden) running Philips version R5.8.1. The 3D-QALAS sequence is not available clinically on Philips version R5.8.1 at the time of this writing, but it is available on Philips version >R12.1.1, therefore the relevant pulse programming environment (PPE) code was backported to version R5.8.1 and compiled into a research patch. In addition, several clinical science keys (CSKs), primarily related to cardiac synchronization and external monitoring devices, had to be overridden in the research patch to run on our MR-Linac system. The relevant acquisition parameters are shown in **Table 1** which were slightly modified from the SyMRI version 15.0.19 recommendations for Philips scanners. This yielded dynamic times of 0, 130.5, 1026, 1922, and 2817 ms.

**Table 1.**
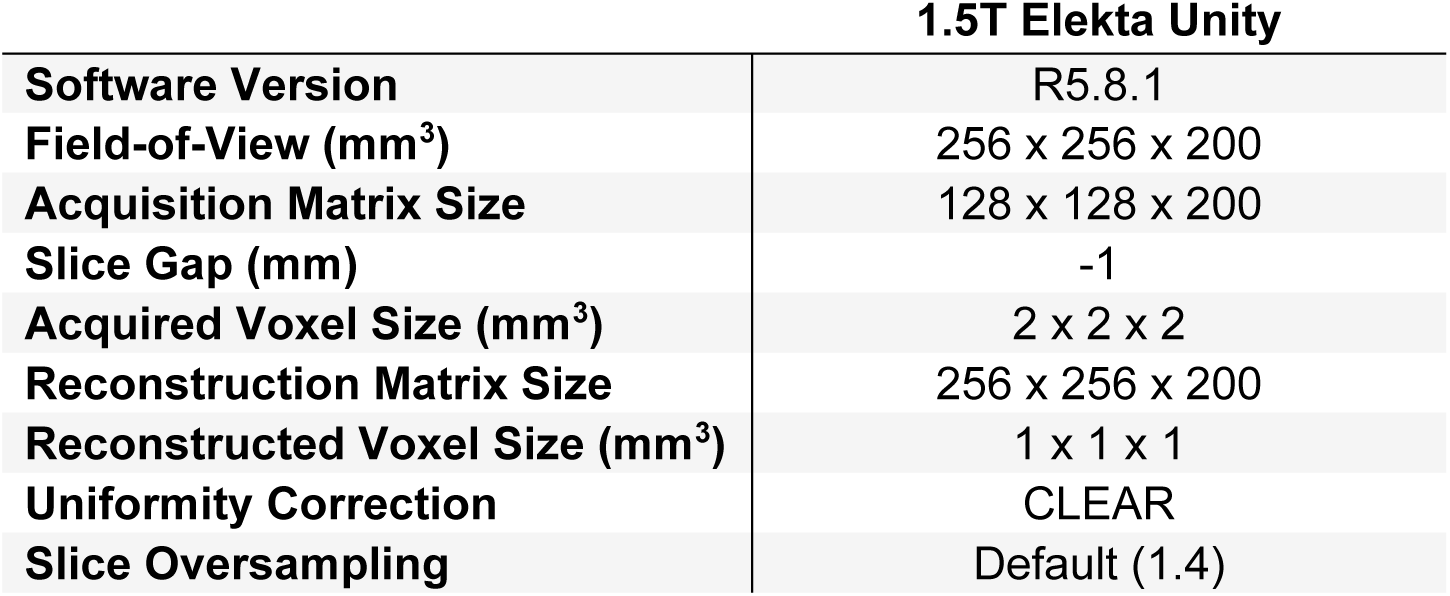
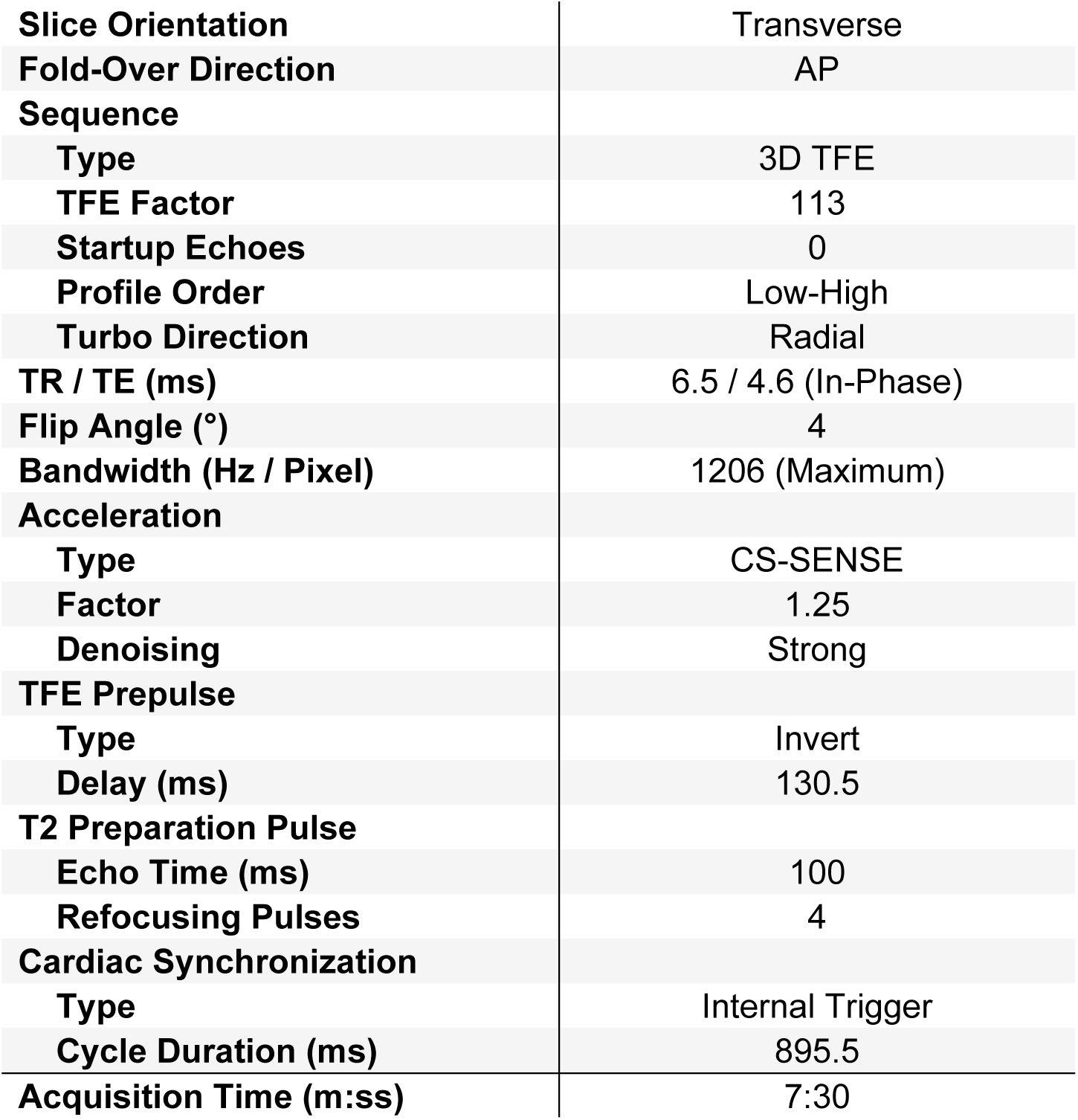
Acquisition parameters across all scanners utilized in this study. Abbreviations: CLEAR = Constant LEvel AppeaRance, AP = anterior to posterior, CS-SENSE = Compressed Sensing (CS) SENSitivity Encoding (SENSE), TFE = turbo field echo, TR = repetition time, TE = echo time.

### 2.2. MRI Acquisition Methodology

Previously developed in collaboration with the National Institute of Standards and Technology (NIST) and the International Society of Magnetic Resonance in Medicine (ISMRM), the CaliberMRI “NIST/ISMRM” Premium System Phantom Model 130 phantom (CaliberMRI; Boulder, CO, USA) was used as a reference for NIST-traceable T1, T2, and PD values and to assess geometric accuracy using the built-in fiducial array^39,40^. The temperature was confirmed to be near 20°C in accordance with the phantom reference values prior to scanning using an analog thermometer placed next to the bore. Since reference data for this phantom was not available at 1.5T at the time of this writing, but only at 3T, we used a previous version of this phantom with both 1.5T and 3T reference data and created a rough conversion factor for quantitative T1, T2, and PD using a linear line-of-best-fit. The resulting conversion factors were 0.93*x + 15.43 ms for quantitative T1, 1.06*x + 26.08 ms for quantitative T2, and 1*x + 0 pu for quantitative PD. To evaluate reproducibility, the 3D-QALAS sequence was acquired across five separate scanning sessions in the NIST/ISMRM phantom and in one session for one healthy volunteer in their 20s. To evaluate repeatability in phantoms within each session, two repetitions were performed per session in a test-retest method^41^.

### 2.3. Data Collection and Image Processing

The resulting magnitude images were processed using the SyntheticMR post-processing software, SyMRI (StandAlone 0.61.0; SyntheticMR AB; Linköping, Sweden) developed for both the 2D-MDME and 3D-QALAS sequences. Since the research patch did not have access to the reconstruction code, it did not correctly assign the temporal position identifier (0020,0100), number of temporal positions (0020,0105), and T2prep time (2005,1712) DICOM tags, so these were modified following image acquisition in accordance with the SyMRI DICOM Conformance Statement using Python 3.12.10.

The geometric accuracy was evaluated using the NIST/ISMRM phantom by determining the coordinates of the center of each vial of the built-in fiducial array and comparing those coordinates to their expected locations^40^ using 3D Slicer^42^ (https://www.slicer.org/). For the phantom analysis, a circular region-of-interest (ROI) was created in each vial across a single slice of the reconstructed quantitative T1 map using 3D Slicer. A margin around the edge was left to account for ringing artifacts and partial volume effects^43^. For the OOI analysis in the healthy volunteer, the white matter, grey matter, and whole brain was automatically contoured using SyMRI and the resulting mean and standard deviation of the quantitative T1, T2, and PD values inside each structure were extracted. SyMRI also produced the estimated white matter, grey matter, cerebral spinal fluid (CSF), myelin, and non-aqueous content maps which further yielded the calculated myelin parenchymal fraction (MyCPF), brain parenchymal fraction (BPF), cerebral spinal fluid (CSF), myelin content (MyC), brain parenchymal volume (BPV), and intracranial volume (ICV). These measured values were compared to references curves derived from a combination of the results from McAllister et al. 2017 for children^44^ and Vågberg et al. 2016 for adults^45^. Additional synthetic T1-weighted, T2-weighted, T2-weighted fluid attenuated inversion recovery (FLAIR), proton density-weighted, phase-sensitive inversion recovery (PSIR), and double inversion recovery (DIR) with white matter suppression maps were generated for qualitative analysis.

### 2.4. Statistical Analysis

All statistical methodology and analytic reporting were formulated using the Statistical Analyses and Methods in the Published Literature (SAMPL) guidelines^46^. Distortion was assessed in each vial by determining the center location of each fiducial, calculating its distance from the cetner fiducial from the image, and comparing these measurements to the nominal distance of 40 mm between orthogonal fiducials. The mean and standard deviation of each vial inside the NIST/ISMRM phantom were extracted and compared to manufacturer reference values. The Lin’s Concordance Correlation Coefficient^47^ (LCCC) was used to evaluate direct agreement to reference values while a Bland-Altman plot^48^ was used to assess bias. In phantoms, the coefficient of variation (CoV) was used to assess repeatability, reproducibility, and noise within the uniform vials of the phantom. For all calculations, analysis was restricted to clinical ranges inside the head and neck^49^ of T1 (200 – 2500 ms), T2 (35 – 120 ms), and PD (20 – 120 pu).

## 3. Results

### 3.1. Phantom Analysis

The measured geometric distortion using the built-in fiducial array of the NIST/ISMRM is shown in **Figure 1**. The distortion at the center was 0 mm since this was used as the reference point. Since the fiducial points were separated by 40 mm, this was the next lowest distance from the center where the median [interquartile range (IQR)] was 1.83 [1.31 – 2.29] mm. The further the fiducials were from the center, the lower the metrics changed with the largest distortion seen at 89.44 mm from the center, the furthest measured point, with a median [IQR] of 1.98 [1.49 – 2.40] mm.

**Figure 1.**
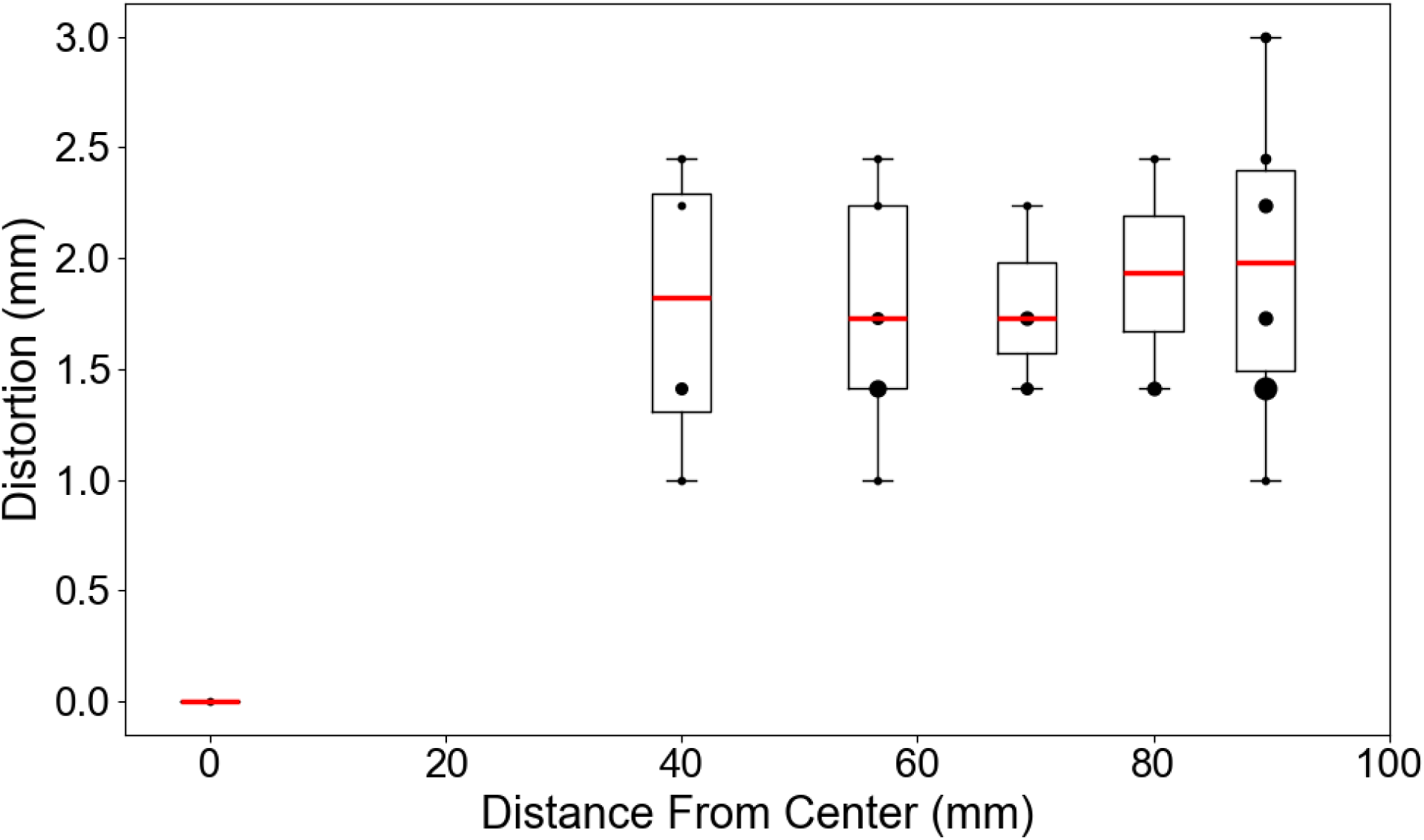
The quantified geometric distortion as a function of distance from the center fiducial point of the NIST/ISMRM phantom in boxplot form with overlayed scatter plot with the dot size representing the number of measurements at that coordinate.

After the geometric distortion was quantified and deemed clinically acceptable, the concordance between the measured quantitative T1, T2, and PD values against the NIST/ISMRM reference values was computed using all ten measurements (five sessions with one test-retest per session) as shown in **Figure 2**. The linear slopes of agreement for the quantitative T1, T2, and PD were 1.02 (LCCC = 1.00), 1.09 (LCCC = 0.90), and 0.99 (LCCC = 1.00), respectively. Both the measured quantitative T1 and PD were strongly linear while the measured quantitative T2 had positive bias against the NIST/ISMRM phantom at both low and high T2 values at the edges of the chosen clinical range of 35 – 120 ms. If the quantitative T1 and PD plots were extended to lower values, then a stabilization of the measured values would be seen, indicating that the chosen minimum clinical range of 200 ms and 20 pu were lower limits for the 3D-QALAS sequence at the current acquisition parameters.

**Figure 2.**
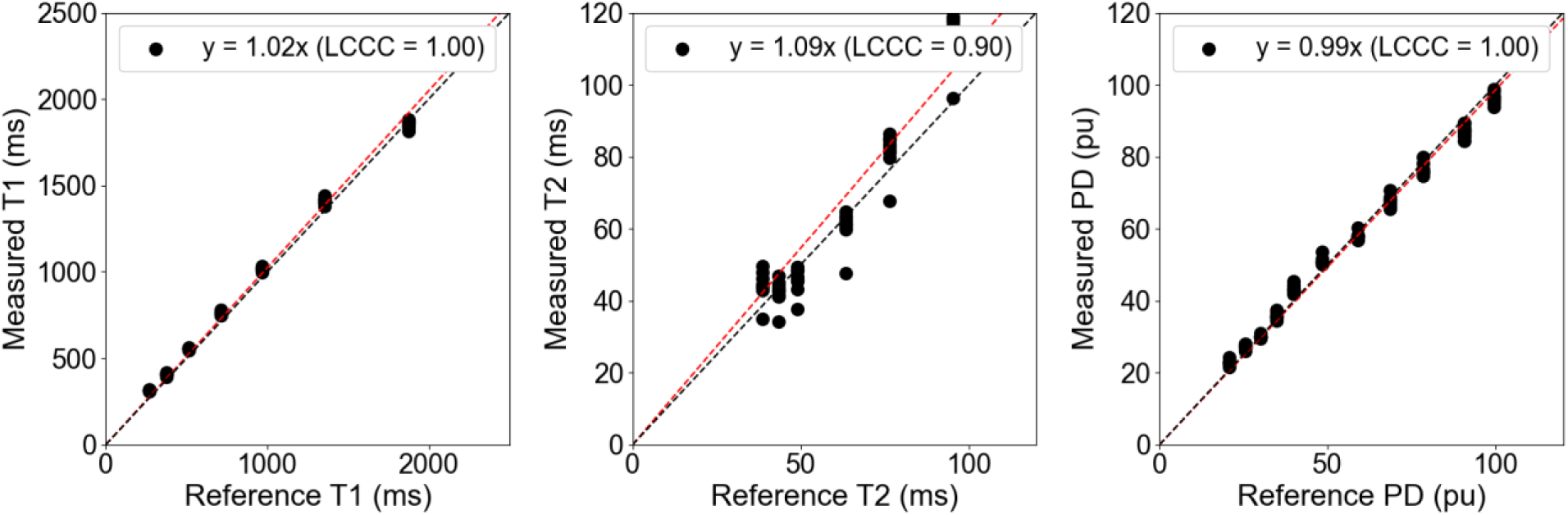
The correlation plot between the measured and reference quantitative T1 (left), T2 (center), and PD (right) across all ten measurements (five sessions with one test-retest per session) of the 3D-QALAS sequence on the 1.5T MR-Linac. Abbreviations: LCCC = Lin’s Concordance Correlation Coefficient.

The corresponding Bland-Altman plots for **Figure 2** is shown in **Figure 3**. All 95% limits of agreement included the 0 value indicating no observed systemic bias across all ten measurements, even though a roughly 50 ms positive bias in the measured quantitative T1 can be seen. Unexpectedly, almost half of the measured quantitative T1 values near 2000 ms exceeded the 95% limits of agreement indicating that this may be the edge of confident quantitative assessment. A similar trend was observed for the quantitative T2 values above 100 ms. Almost all of the measured quantitative PD values remained within the 95% limits of agreement which was centered around 0 pu with a range of 5 ms in both the positive and negative bias directions. The corresponding ranges for the quantitative T1 and T2 was 50 ms and 20 ms in either direction, respectively.

**Figure 3.**
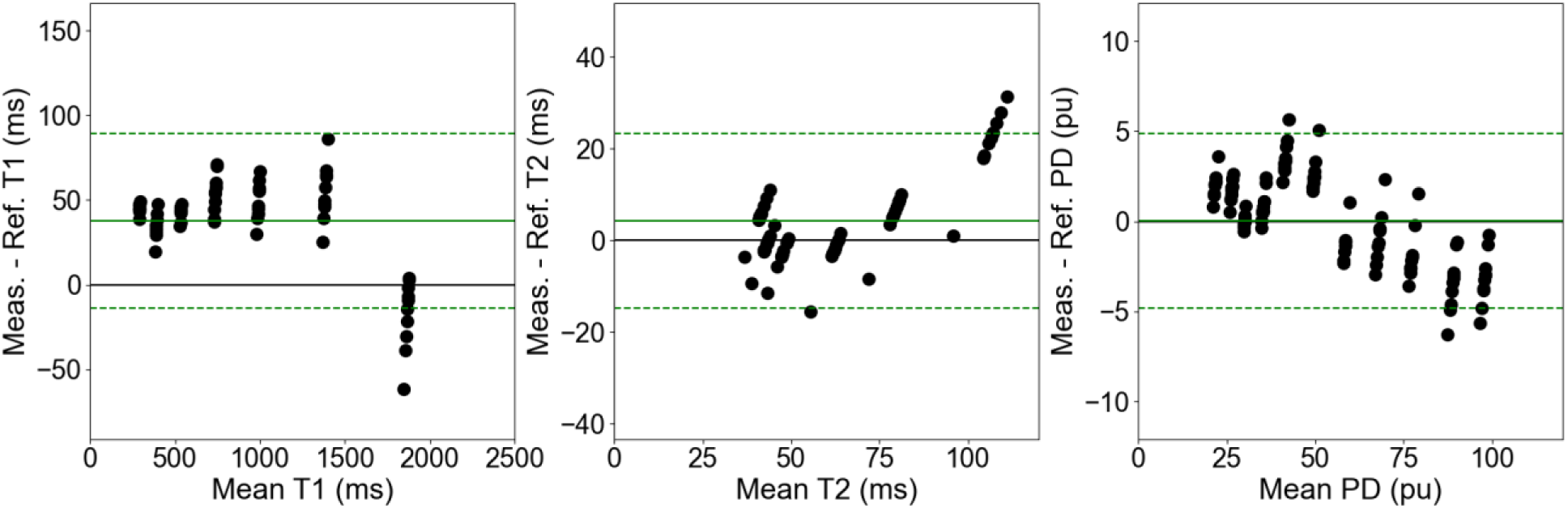
The Bland-Altman plot between the measured and reference quantitative T1 (left), T2 (center), and PD (right) across all ten measurements (five sessions with one test-retest per session) of the 3D-QALAS sequence on the 1.5T MR-Linac. The solid green line represents the mean across all biases while the dashed green lines represent the 95% limits of agreement.

The corresponding repeatability of the two repetitions averaged across the five sessions and the reproducibility of the five sessions averaged across the two repetitions for the measured quantitative T1, T2, and PD is shown in **Figure 4**. Both the repeatability and reproducibility was under 2% for the quantitative T1 values while being under 3% for the quantitative PD values and under 8% for the quantitative T2 values. For all vials tested, the reproducibility remained higher than the repeatability. This difference was minimized for the quantitative T1 values where they were nearly identical and maximized for the quantitative PD values where the reproducibility was nearly twice as high. The quantitative T2 values fell within these two trends.

**Figure 4.**
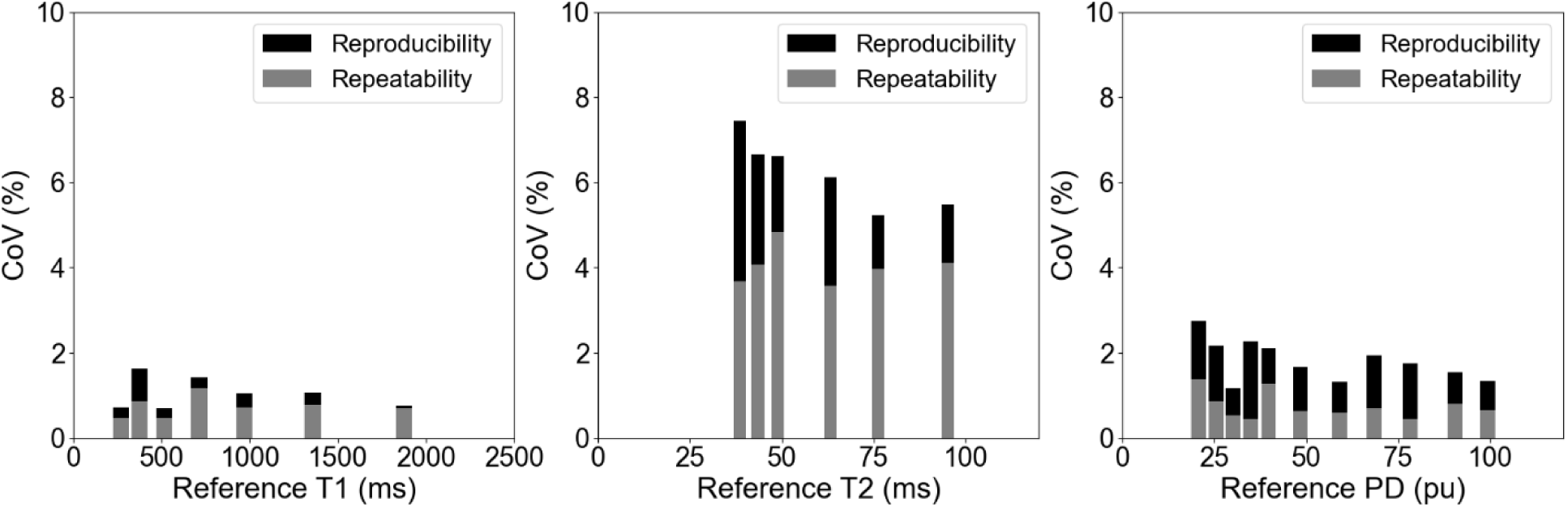
The repeatability of the two repetitions averaged across the five sessions and the reproducibility of the five sessions averaged across the two repetitions for the quantitative T1 (left), T2 (center), and PD (right) from the 3D-QALAS sequence on the 1.5T MR-Linac as measured by the coefficient-of-variation (CoV).

### 3.2. Healthy Volunteer Analysis

The quantitative T1, T2, and PD maps in axial, coronal, and sagittal orientations in the healthy volunteer are shown in **Figure 5**. The primary purpose of this figure was to demonstrate the benefits of the isotropic spatial resolution on different viewing orientations without the blurry appearance seen in 2D-MDME which traditionally has thick (∼3-4 mm) slices. This healthy volunteer had a permanent retainer which led to the signal void in the oral cavity for all images shown for this volunteer. Slightly higher image noise can be seen in the quantitative T2 maps which was reflected in the previously shown concordance, repeatability, and reproducibility plots where is performed the worst compared to the quantitative T1 and PD.

**Figure 5.**
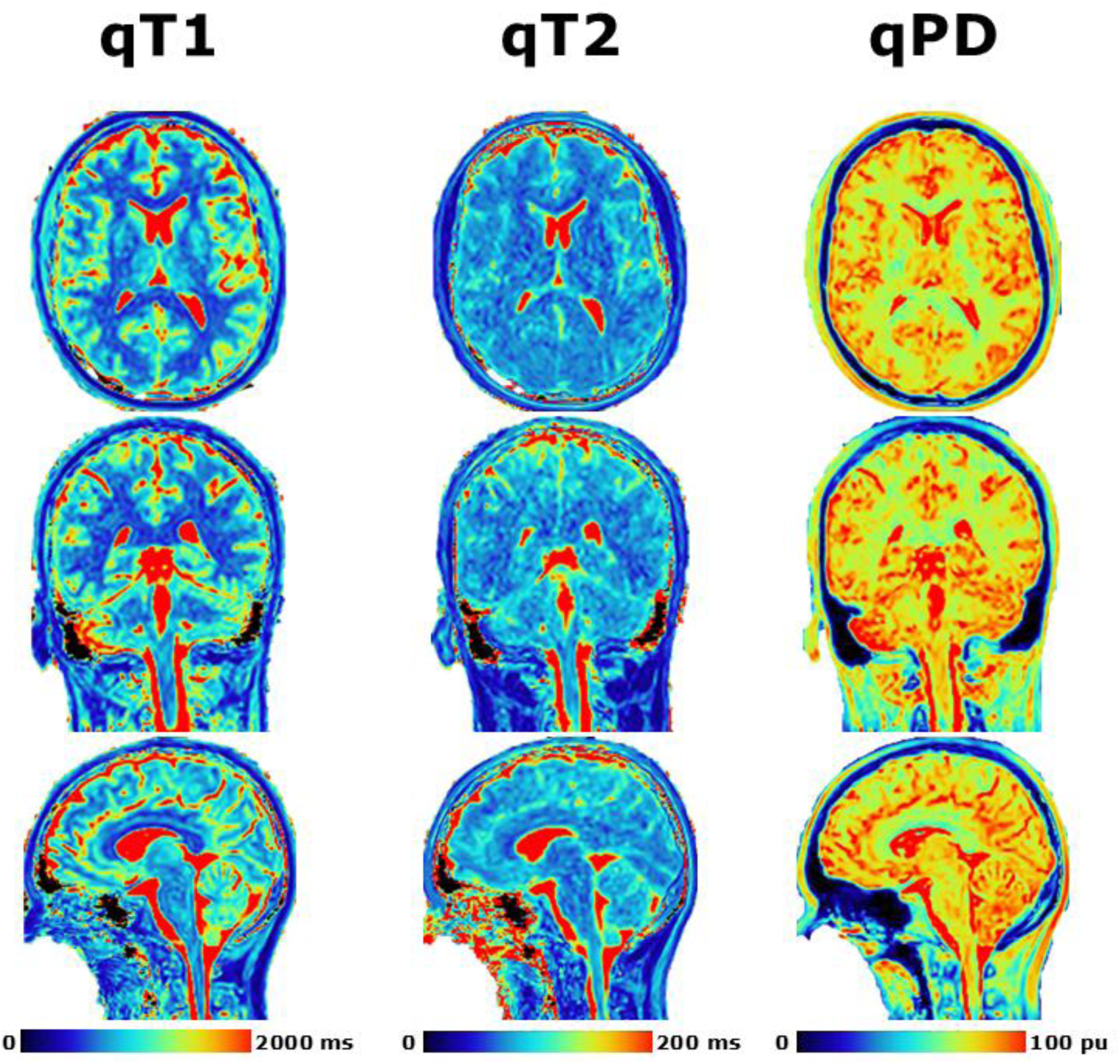
Demonstration of post-processing available from SyMRI combined with 3D-QALAS at high isotropic resolution in a healthy volunteer. Shown are the quantitative T1 (left), quantitative T2 (middle), and quantitative PD (right) in the axial (top), coronal (middle), and sagittal (bottom) orientations.

The quantitative results from **Figure 5** for the automatically delineated white matter, grey matter, and whole brain are shown in **Table 2**. The white matter had decreased mean T1 of 725 ms compared to 1183 ms in grey matter while the T2 was only slightly lower at 76 ms compared to 88 ms and the PD was lower at 71.9 pu compared to 82.8 pu. The white matter represented 34% of the whole brain while grey matter was 57% with the remaining content being CSF and other macromolecules not captured within the same quantitative ranges as white matter and grey matter.

**Table 2.** The calculated total structure volume and mean (standard deviation) of the quantitative T1, T2, and PD values inside the white matter, grey matter, and whole brain.

|  | White Matter | Grey Matter | Whole Brain |
| --- | --- | --- | --- |
| Volume (ml) | 429.1 | 719.2 | 1256.8 |
| T1 (ms) | 725 (128) | 1183 (351) | 1025 (381) |
| T2 (ms) | 76 (10) | 88 (20) | 86 (25) |
| PD (pu) | 71.9 (4.7) | 82.8 (7.5) | 78.5 (8.8) |

The synthetic T1-weighted, T2-weighted, T2-weighted fluid attenuated inversion recovery (FLAIR), proton density-weighted, phase-sensitive inversion recovery (PSIR), and double inversion recovery (DIR) with white matter suppression images in axial, coronal, and sagittal orientations are shown in **Figure 6**. The T1-weighted images appeared to have the lowest amount of noise with the most noise seen in the T2-weighted images. Adding the FLAIR component led to good CSF suppression with a very small amount of residual high intensity regions where CSF is expected. Only a moderate amount of noise was seen in the PD-weighted images while effectively providing an average of the T1-weighted and T2-weighted images.

**Figure 6.**
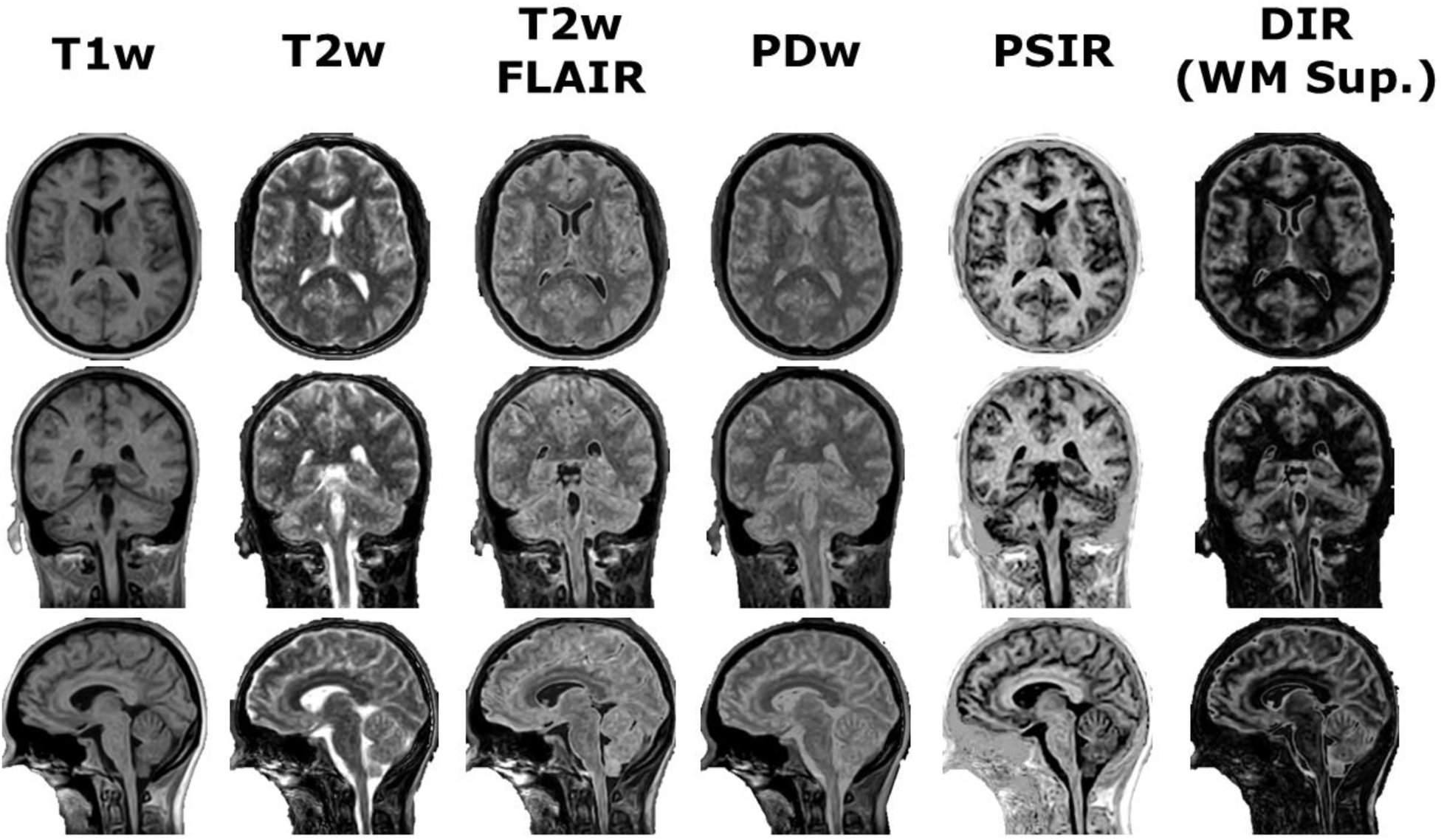
Demonstration of post-processing available from SyMRI combined with 3D-QALAS at high isotropic resolution in a healthy volunteer. Shown from left to right are the T1-weighted, T2-weighted, T2-weighted fluid attenuated inversion recovery (FLAIR), proton density-weighted, phase-sensitive inversion recovery (PSIR), and double inversion recovery (DIR) with white matter suppression synthetic images in the axial (top), coronal (middle), and sagittal (bottom) orientations.

Finally, moderate noise was seen in the PSIR images while providing a unique contrast for structure delineation while minimal noise was seen in the DIR images with mostly effective white matter suppression.

The calculated white matter, grey matter, cerebrospinal fluid, myelin, and non-aqueous component synthetic images in axial, coronal, and sagittal orientations overlayed onto the synthetic T1-weighted image is shown in **Figure 7**. Both white matter and grey matter appear in the correct locations of the brain, however some decreased confidence (i.e., a considerable number of voxels are below 100%) is seen particularly in the superior port of the images. The CSF map provides a clear distinction around both the sulci and gyri as well as the ventricle with high confidence. Similar to the white and grey matter, the myelin images have decreased confidence throughout the image due to the noise in the quantitative T2 maps. This noise is also generally reflected in the non-aqueous content image as well.

**Figure 7.**
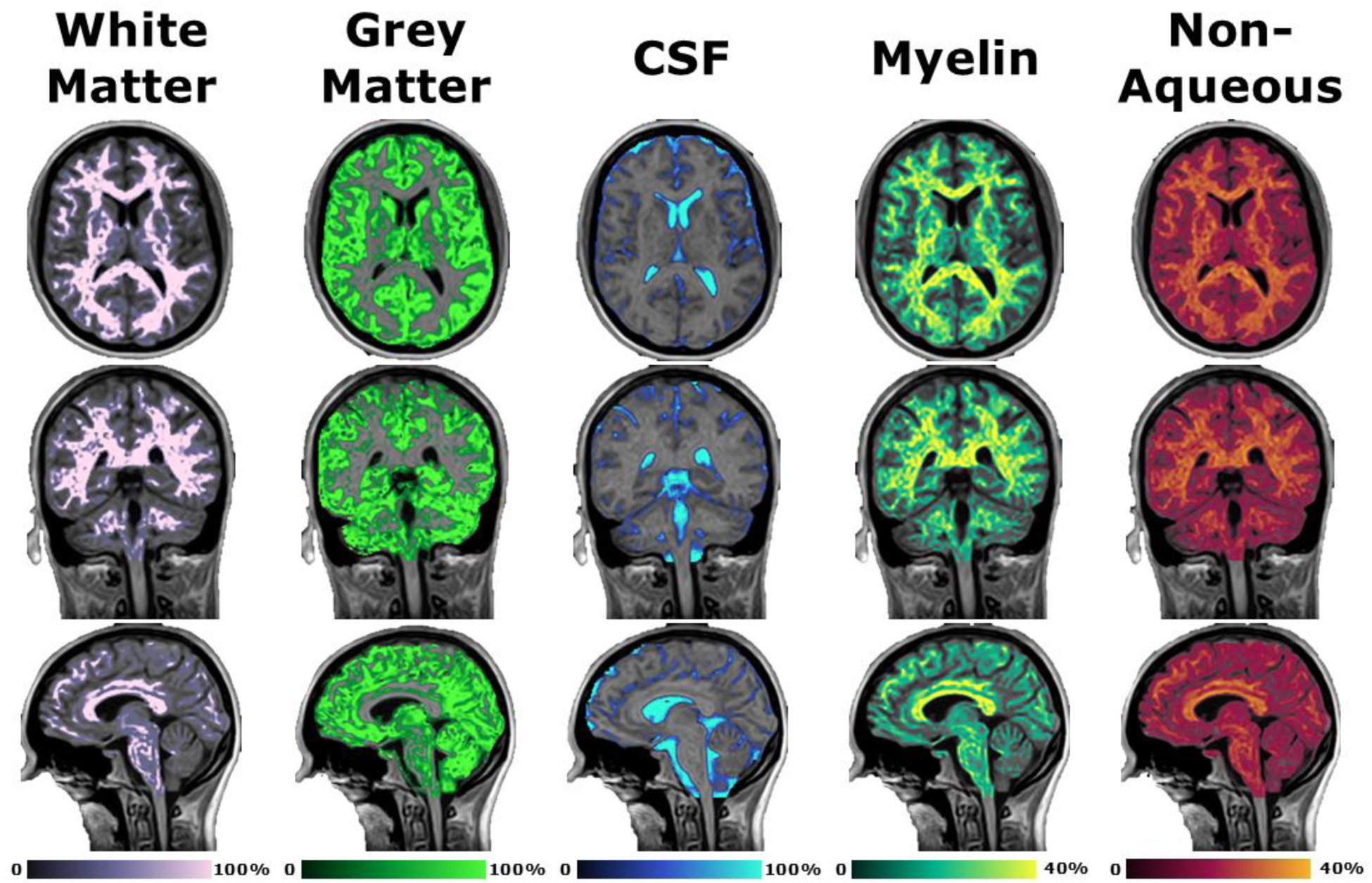
Demonstration of post-processing available from SyMRI combined with 3D-QALAS at high isotropic resolution in a healthy volunteer. Shown from left to right are the white matter, grey matter, cerebrospinal fluid, myelin, and non-aqueous component synthetic images in the axial (top), coronal (middle), and sagittal (bottom) orientations overlayed onto the synthetic T1-weighted image.

A visualization of the measured MyCPF, BPF, CSF, MyC, BPV, and ICV values in the healthy volunteer in their 20s compared to age-dependent reference values are shown in **Figure 8**. The measured value across all six metrics was within the reference range for healthy individuals in their 20s. Quantitatively, the MyCPF was 12.9%, the BPF was 90.9%, the CSF was 125.5 ml, the MyC was 161.8 ml, the BPV was 1256.8 ml, and the ICV was 1382.2 ml. In addition to these reported values, the total white matter volume was 429.1 ml, the total grey matter volume was 719.2 ml, and the total non-aqueous content was 230.5 ml.

**Figure 8.**
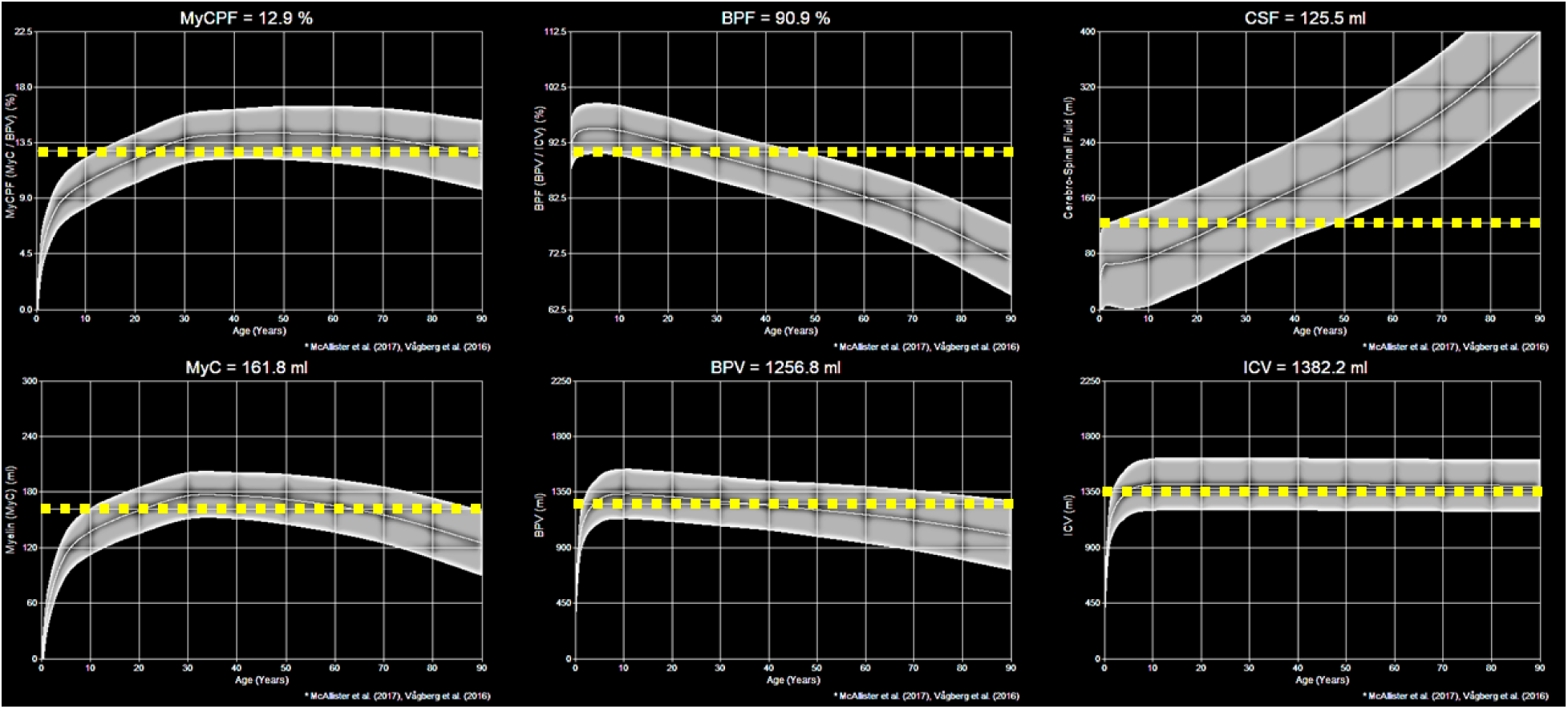
Demonstration of post-processing available from SyMRI combined with 3D-QALAS at high isotropic resolution in a healthy volunteer. Shown are the myelin parenchymal fraction (MyCPF), brain parenchymal fraction (BPF), cerebral spinal fluid (CSF), myelin content (MyC), brain parenchymal volume (BPV), and intracranial volume (ICV) in the healthy volunteer in their 20s. The dashed yellow line is the measured content while the grey bands are the predicted reference values for healthy individuals.

## 4. Discussion

### 4.1. Summary

In this study, we demonstrated the technical feasibility of the 3D-QALAS acquisition on the 1.5T MR-Linac by assessing its geometric accuracy, concordance, repeatability, and reproducibility in both phantoms and a healthy volunteer. The slope and LCCC between the measured and phantom reference values was 1.02 (LCCC = 1.00), 1.09 (LCCC = 0.90), and 0.99 (LCCC = 1.00) for the quantitative T1, T2, and PD values, respectively, while the median distortion remained below 2 mm, or under two voxels. The repeatability and reproducibility coefficient-of-variation (CoV) was under 5% and 8%, respectively, for all measured values. The measured brain volumes in the healthy volunteer was within expected age-adjusted reference values.

### 4.2. Limitations and Future Work

The method to determine geometric distortion had limitations of precision due to 3D Slicer only allowing segmentations to be drawn on finite voxels, not partial voxels. Therefore, after a 0 mm distortion, the next lowest distortion would be 1 mm due to the 1 mm voxel resolution of our 3D-QALAS acquisition. Therefore, a median geometric distortion of between 1 – 2 mm across the entire fiducial array should be acceptable for most applications since it is only at the order of 1 – 2 voxels. Future work should be done to acquire 3D-QALAS at higher spatial resolutions to improve this precision or use a software that allows partial voxel segmentations. Direct reproducibility across multiple 1.5T MR-Linac machines could not be determined since our institution only has one, therefore further work should be done to confirm the results shown here which was done by test-retest across multiple independent sessions.

Although efficient compared to traditional quantitative relaxometry mapping techniques, a total acquisition time of over seven minutes is higher than desired and should be reduced in further studies. A current hinderance to further reducing the acquisition time is the compressed sensing (CS-SENSE) factor which would lead to the introduction of image noise^50^. However, combining 3D-QALAS with Wave–Controlled Aliasing In Parallel Imaging (Wave-CAIPI^51^) readouts can reduce and unify g-factors^52^, especially when combined with model-based deep learning (Wave-MoDL^53^) and zero-shot deep learning subspaces (Zero-DeepSub^54^). Another approach is to use offline compressed sensing reconstruction using the raw image data which has been previously demonstrated successfully on the 1.5T MR-Linac for quantitative T1 mapping using non-Cartesian acquisitions^55^.

Due to these concerns, future work should focus on optimization of the acquisition parameters^56^ to better quantify relaxometric properties for tissues in the head and neck, preferably in an R-IDEAL Stage 2a study. Some acquisition parameters that should be focused on during the optimization would be the TFE inversion prepulse delay time which would shift the location of the first readout following the inversion pulse primarily affecting the T1 quantification, the T2-preparation delay time which would affect the T2 quantification, and the internal trigger delay time which would adjust the spacing between readouts primarily affecting the T1 quantification. These optimizations could also minimize the inhomogeneities seen in the quantitative T2 maps in **Figure 1**. Further optimization could then be made to the CS-SENSE factor and its denoising factor for reduced acquisition times, larger field-of-views, and increased signal-to-noise ratio (SNR) – especially since the water-fat shift (WFS) must be minimum to minimize chemical shift induced signal intensity biases^57^.

### 4.3. Extended Applications

Building on the work presented here investigating 3D-QALAS, this high resolution isotropic method to quantitatively map multiparametric relaxometric properties can be applied synergistically with recent efforts to define probabilistic tumor spread^58^. This approach, in conjunction with the high temporal resolution of the MR-Linac, can bring new insights into how tumors and OOIs respond to radiation therapy allowing for advanced tumor control probability (TCP^59^) and normal tissue complication probability (NTCP^60^) modeling. Currently, the 3D-QALAS sequence utilizes a one echo readout, however multiple echoes can be added which would allow for quantitative T2* mapping and it’s relative derived maps such as in the Multiple-echoes with Optimized Simultaneous Acquisition (MIMOSA) technique^61^. Another approach is to add an additional T2-preparation module which can be employed to more accurately quantify the myelin water fraction (MWF) as demonstrated in the MWF-QALAS^62^ and Omni-QALAS^63^ techniques. Another interesting future application of 3D-QALAS on the MR-Linac is the reconstruction of time-of-flight MR angiography images using deep learning^64^ which can be used for improved target planning^65–67^. Finally, the generation of synthetic computed tomography (CT) images^68^ at 1 mm isotropic resolution derived from the three quantitative maps from 3D-QALAS would help MR-only workflows^69–71^, reducing burden on patients while minimizing delay in care. Integration of these advanced, high resolution, quantitative MRI techniques into the adaptive head and neck radiation workflow has the potential to help guide the reduction of target margins^72^, especially when combined with prospective motion correction techniques^73^.

## 5. Conclusion

The 3D-QALAS sequence is a promising approach to acquire multiparametric quantitative MRI relaxometric maps and synthetically generated contrast-weighted maps at high resolution isotropic voxel sizes. In this paper, we demonstrated the potential of 3D-QALAS to transform the quantitative image biomarker capacity of the 1.5T MR-Linac in the following ways: 1) achievement of quantitative T1 / T2 / PD maps at 1 mm isotropic voxel size in under eight minutes, 2) derivation of pre-registered synthetic contrast-weighted images at equivalent spatial resolution, and 3) repeatability under 5%, reproducibility under 8%, and median spatial accuracy under 2 mm. With this technique, a new era is emerging where quantitative MRI maps can be acquired at the same spatial resolution as traditional anatomical imaging (i.e., T2-weighted), enabling advanced tumor and OOI quantification and characterization, ideal for adaptive radiation therapy applications on the MR-Linac.

